# Proportion of paediatric admissions with any stage of noma at the Anka General Hospital, northwest Nigeria

**DOI:** 10.1101/2023.07.10.23292440

**Authors:** Elise Sarah Farley, Miriam Njoki Karinja, Abdulhakeem Mohammed Lawal, Michael Olaleye, Sadiya Muhammad, Maryam Umar, Fatima Khalid Gaya, Shirley Chioma Mbaeri, Mark Sherlock, Deogracia Wa Kabila, Miriam Peters, Joseph Samuel, Guy Maloba, Rabi Usman, Saskia van der Kam, Koert Ritmeijer, Cono Ariti, Mohana Amirtharajah, Grégoire Falq

## Abstract

**Introduction:** Noma is a rapidly spreading infection of the oral cavity which mainly affects young children, and without early treatment, can have a high mortality rate. Simple gingivitis is a warning sign for noma, and acute necrotizing gingivitis is the first stage of noma. The epidemiology of noma is not well understood. We aimed to gather evidence on the epidemiology of noma and its association with malnutrition for use in program planning.

**Methods:** We conducted a prospective observational study enrolling patients aged 0 to 12 years who were admitted to the Anka General Hospital, Zamfara, northwest Nigeria. Consenting caregivers of participants were interviewed at admission. Participants had anthropometric and oral exams at admission and discharge.

**Findings:** Of the 2346 participants, 58 (2.5%) were diagnosed with simple gingivitis and six (n=0.3%) with acute necrotizing gingivitis upon admission. Of those admitted to the Inpatient Therapeutic Feeding Centre (ITFC), 3.4% (n=37, CI 2.5 - 4.7%) were diagnosed with simple gingivitis upon admission compared to 1.7% of those not admitted to the ITFC (n=21, CI 1.1 - 2.6%) (p=0.008). Risk factors identified for having simple gingivitis include being aged over two years (2 to 6yrs old, odds ratio (OR) 3.4, CI 1.77 - 6.5; 7 to 12 yrs OR 5.0, CI 1.7 - 14.6; p=<0.001) and being admitted to the ITFC (OR 2.1; CI 1.22 -3.62).

**Conclusion:** Our study showed a small proportion of those admitted to the Anka General Hospital had simple or acute necrotizing gingivitis. Those admitted to the ITFC were more likely to have simple gingivitis. The lack of access to and uptake of oral health care indicates a strong need for oral exams to be included in routine health services. This provision could improve the oral status of the population and decrease the chance of patients developing noma.

**Author summary:** Noma is a rapidly spreading infection of the oral cavity which mainly affects young children and without early treatment, can have a high mortality rate. Simple gingivitis is a warning sign for noma, and acute necrotizing gingivitis is the first stage of noma. We aimed to gather evidence on the epidemiology of noma and its association with malnutrition by conducting a prospective observational study enrolling 2346 patients aged 0 to 12 years who were admitted to the Anka General Hospital, Zamfara, northwest Nigeria. Consenting caregivers of participants were interviewed at admission. Participants had anthropometric and oral exams at admission and discharge. Our study showed a small proportion of those admitted to the Anka General Hospital had simple or acute necrotizing gingivitis. Those admitted to the ITFC were more likely to have simple gingivitis. The lack of access to and uptake of oral health care indicates a strong need for oral exams to be included in routine health services. This provision could improve the oral status of the population and decrease the chance of patients developing noma.

## Introduction

Noma is an infection of the oral cavity which can cause the disintegration of the cheek, nose and/or eye in a few weeks [1]. If untreated in the early reversible stages, noma has a reported 90% mortality rate [2] and mainly affects children aged between two and six years [3]. Deaths in noma patients are primarily due to starvation, aspiration pneumonia, respiratory insufficiency or sepsis [4,5]. With timely antibiotic treatment, wound debridement and nutritional support, morbidity and mortality from noma greatly decrease [1]. For those who survive, noma often leads to severe facial disfigurement, functional issues and stigmatization [1,2,6,7]. The WHO classifies noma into stages: Warning Sign: simple gingivitis, Stage 1: acute necrotizing gingivitis, Stage 2: oedema, Stage 3: gangrene, Stage 4: scarring and Stage 5: sequelae [2].

There is limited research on noma and as a result, many gaps in knowledge exist around the disease [1]. The aetiology of noma is unknown but thought to be multifactorial [1]. Risk factors for noma include poverty, poor oral hygiene, poor access to routine childhood vaccinations, limited access to quality health care and immunosuppression resulting from comorbidities such as malnutrition, measles and HIV [1,8–16]. Malnutrition is frequently listed as a risk factor for noma, however, evidence supporting this theory is largely based on case reports and case series [11,17–27] and a handful of primary studies [8–10,28–30]. The epidemiology of the disease is also not well understood. The WHO estimates that 140, 000 children contract noma each year globally [31]. In 2003, a northwest Nigerian study estimated the noma incidence was 6.4 per 1000 children from 1996 to 2001 [32]. A recent study estimated that the period prevalence of noma from 2010-2018 was 1.6 per 100,000 population at risk in northern Nigeria [33]. A 2019 prevalence study in Sokoto and Kebbi states, Nigeria, identified that in children aged 0 to 15 years (n=7122), simple gingivitis was diagnosed in 3.1% (n=181; 95% confidence interval (CI) 2.6-3.8), acute necrotizing gingivitis in 0.1% (n=10; CI 0.1-0.3), and oedema in 0.05% (n=3; CI 0.02-0.2), no late stage noma cases were identified [34].

We conducted a prospective observational study assessing the proportion of pediatric admissions at the Anka General Hospital with any stage of noma, to add to the understanding of the epidemiology of the disease and examine the links between noma and malnutrition.

## Methods

### Study design and data collection

From 1^st^ June to 24^th^ October 2021, we conducted a prospective observational study utilising face-to-face interviews with caregivers of paediatric patients (aged under 12 years) admitted to the Anka General Hospital, northwest Zamfara, Nigeria and physical examinations of the patients themselves. All caregivers of patients admitted between 09h00 and 16h00 Monday to Sunday were approached on the day of admission by the study nurse. Consenting individuals were enrolled (discussed in the Ethics section below). A tablet-based RedCap data collection form was used to collect the study specific data on admission (sociodemographic characteristics, infant and current feeding practices, oral health practices, health care access, oral examination, mid upper arm circumference, weight, and height) and at discharge (oral exam and access to routinely collected medical records).

### Sample size

We aimed to have a sample size large enough to estimate the proportion of paediatric admissions with any stage of noma admitted to the Inpatient Therapeutic Feeding Centre (ITFC) or other wards. This calculation was based on the following assumptions:

- 6%- Inpatient department proportion of patients diagnosed with any stage of noma (as this had a lower proportion of any stage of noma diagnosed than the ITFC [35])
- 95% confidence interval
- 1.5% margin of error (i.e., can estimate a proportion between 4.5 and 7.5% assuming a true proportion of 6%)

The sample size calculation showed that it was necessary to include 963 patients in the ITFC and 963 patients in the other wards.

### Analysis

We performed a descriptive analysis of the data collected. Categorical variables are shown as frequencies and percentages. Continuous variables were summarised using means and standard deviations (SD) or medians and interquartile ranges (IQR) depending on normality. Missing data numbers are recorded in each table.

Age was categorised into 0-1 years, 2-6 years and 7-12 years. This age categorisation was selected as children aged between two and six years have previously been reported to be at higher risk of noma [3].

Malnutrition estimates are not reported for participants aged zero to five months as we did not have access to the confirmed birthweights of these children [36].

For children aged six months to 5 years, using Mid-Upper Arm Circumference (MUAC) measurements taken upon admission, we estimated the proportion of severe acute malnutrition (SAM, MUAC <115mm), moderate acute malnutrition (MAM, between MUAC ≥115 and <125mm) and global acute malnutrition (GAM, MUAC <125mm) [36].

For those aged six to 12 years, we report the WHO recommended Body Mass Index (BMI)-for-age Z-scores [36]. BMI was calculated based on the weight (in kg) divided by the square of the height (in m) of the individual. BMI-for- age are presented as Z-scores based on the 2007 WHO Growth Reference [36].

Wealth scores were calculated by assigning a value of one to each of the following items owned by the family: cell phone, radio, motorbike, tractor, bicycle, car (these items were chosen based on consultation with local researchers, knowledgeable about the context). The minimum wealth score was zero and the maximum was six, all items weighed equally in the scoring.

We compared the proportion of admissions with each stage of noma between those admitted to the ITFC and those admitted to other wards using a Pearson’s chi-square test.

We calculated the number of admissions who had any stage of noma upon admission, and the outcome of those patients (noma stage at discharge) for all those who had oral examinations on both admission and discharge.

We assessed risk factors for simple gingivitis using univariate logistic regression (patient age, wealth score, ITFC admission status, if the child was sick during the past 3 months, parent primary caretaker, if child had been given colostrum at birth, if the child eats pap, if the child was vaccinated, SAM, MAM and GAM status upon admission, measles, and malaria diagnoses). These variables were selected as they are reported risk factors for noma [1,8–27]. We conducted an exploratory analysis of risk factors for acute necrotizing gingivitis; proportions are reported.

All data analysis was conducted with Stata 17 (StataCorp LP, College Station, TX, USA).

### Ethics

The Médecins Sans Frontières Ethics Review Board (ERB) (2017), Nigerian Federal Ministry of Health ERB (NHREC/01/01/2007-29/04/2021), Zamfara Ministry of Health ERB (ZSHREC01112020) and the Usman Danfodiyo University Teaching Hospital Health Research and Ethics Committee in Nigeria (NHREC/30/012/2019) approved the study protocol.

Caregivers of participants were asked to sign an informed consent form and participants aged between seven and 12 years were informed about the study and asked to sign an assent form after reading/ hearing the information provided on the information sheet. For participants or caregivers who were illiterate, we requested them to provide an initial and a thumb print as a representation of consent, with additional participation from an independent literate witness who checked that the participant/ caregiver understood the contents of the consent form. This witness co-signed the consent form. The information sheet and the informed consent and assent forms were written in English and Hausa.

## Results

### Sociodemographic characteristics

Of those caregivers approached (n=2356), six did not provide consent and four did not provide assent to be part of the study; thus 2346 participants were enrolled and included in the analysis, 2340 of these had a discharge assessment. Caregivers were mainly aged between 18 and 25 years (n=1036, 44.3%) and 26 to 35 years (n=946, 40.5%), female (n=2318, 99.2%) and the mother of the child (n=2211, 94.6%). The main source of income of most families was agriculture (n=1285, 55.0%). Most (n=1696, 72.3%) families owned zero to two items on the wealth score list. Half the child participants were female (n=1181, 50.6%), most were aged between zero and one years (n=1097, 47.1%) or two to six years (n=1135, 48.8%) (Table 1).

**Table 1.** Sociodemographic characteristics, noma study Anka General Hospital 2021

| Caretaker age | N=2346 | % |
| --- | --- | --- |
| 18 - 25 years | 1036 | 44.3% |
| 26 - 35 years | 946 | 40.5% |
| 36 - 45 years | 283 | 12.1% |
| > 46 years | 71 | 3.1% |
| Missing | 10 |  |
| Caretaker sex |  |  |
| Female | 2318 | 99.2% |
| Male | 19 | 0.8% |
| Missing | 9 |  |
| Caretaker education |  |  |
| None | 732 | 31.3% |
| Arabic studies | 1499 | 64.1% |
| Primary school | 54 | 2.3% |
| Secondary school | 50 | 2.1% |
| University | 2 | 0.1% |
| Missing | 9 |  |
| Income |  |  |
| Agriculture | 1285 | 55.0% |
| Trader | 745 | 31.9% |
| Mining | 88 | 3.8% |
| Daily labourer | 53 | 2.3% |
| Civil servant | 43 | 1.8% |
| Livestock | 37 | 1.6% |
| Other | 85 | 3.6% |
| Missing | 10 |  |
| Relationship to child |  |  |
| Mother | 2211 | 94.6% |
| Other | 125 | 5.4% |
| Missing | 10 |  |
| Wealth score (own: cell phone, radio, motorbike, tractor, bicycle, car) |  |  |
| 0-2 items | 1696 | 72.3% |
| 3-6 items | 650 | 27.7% |
| Child gender |  |  |
| Female | 1181 | 50.6% |
| Male | 1154 | 49.4% |
| Missing | 11 |  |
| Child age (years) |  |  |
| 0-1 | 1097 | 47.1% |
| 2-6 | 1135 | 48.8% |
| 7-12 | 95 | 4.1% |
| Missing | 19 |  |

### Health care access and primary diagnoses

Caregivers reported several challenges accessing care including difficulties with transport (n=454, 19.4%) and the hospital being far from their home (n=432, 18.4%). Half of the respondents (n=1176, 50.4%) reported that the child had been sick in the prior three months (not including the illness that made them seek care on the day of enrolment). Of these, almost all (n=1069, 90.9%) sought care for that illness, many from a street pharmacist (n=424, 39.7%) (Table 2).

**Table 2.** Health care access noma study Anka General Hospital 2021

| Challenges when accessing care | N= 2346 | % |
| --- | --- | --- |
| Transport difficult | 454 | 19.4% |
| Far to get to hospital | 432 | 18.4% |
| Difficult to leave other children | 22 | 0.9% |
| Takes a long time to get to hospital | 52 | 2.2% |
| Child very sick, difficult to travel | 6 | 0.3% |
| Difficult to get permission to travel from family | 6 | 0.3% |
| Cost of care too expensive | 13 | 0.6% |
| No challenges | 1505 | 64.2% |
| Has your child been sick in the past 3 month? (other than the sickness that made you seek care today) |  |  |
| No | 1157 | 49.6% |
| Yes | 1176 | 50.4% |
| Missing | 13 |  |
| If yes, did you seek care due to that sickness? (N=1176) |  |  |
| No | 104 | 8.8% |
| Yes | 1069 | 90.9% |
| Where did you seek care? (N=1069) |  |  |
| Hospital | 297 | 27.8% |
| Clinic | 327 | 30.6% |
| Traditional healer | 62 | 5.8% |
| Street chemist, pharmacist | 424 | 39.7% |

Of those respondents who had teeth (n=1907, 81.7%), almost all (n= 1652, 86.6%) cleaned their teeth, mainly using water (n= 1399, 84.7%). The main reasons for children cleaning their teeth were to get clean, bright teeth (n=537, 32.5%) and to get rid of foul breath (n=944, 57.1%). The main reasons for children not cleaning their teeth (n=251), were not knowing the benefits of cleaning (n=18, 7.2%) and forgetting to clean their teeth (n= 19, 7.6%). Most children (n=2069, 88.5%) had not had any dental issues in the three months prior to study enrolment. The most common oral health issues reported (n=265, 11.4%) were a sore in the mouth (n=149, 56.2%), bleeding gums when touched (n=88, 33.2%) and sore gums (n=34, 12.8%). Almost all the respondents (n=2274, 97.4) had not had an oral health check in the past year (Table 3).

**Table 3.**
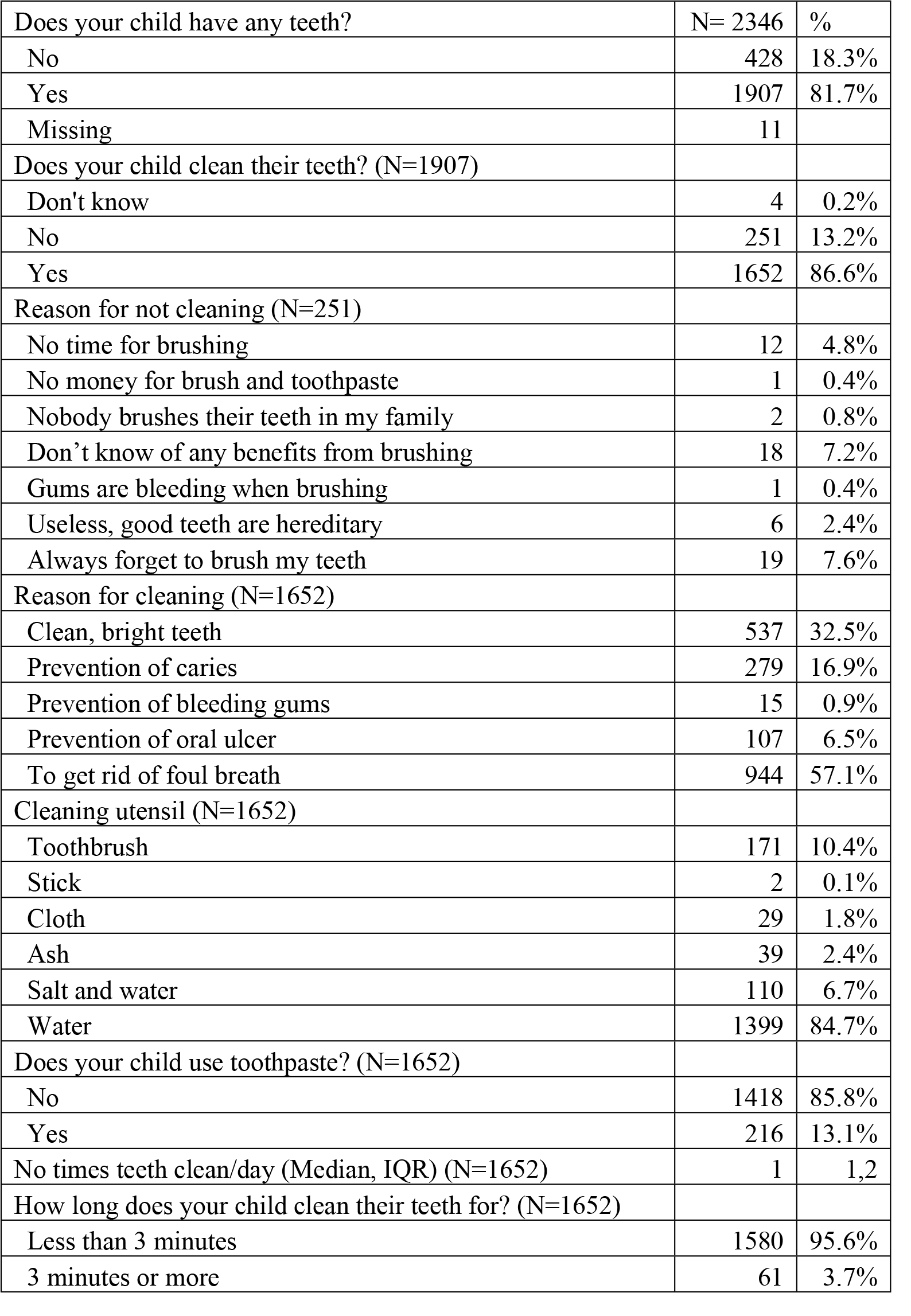
Oral hygiene and health care noma study Anka General Hospital 2021

The most common primary diagnosis of patients admitted to the hospital were malaria (n= 1086, 46.3%) and severe acute malnutrition (n=977, 41.6%). The median number of days patients spent in the hospital was three (IQR 2, 5) (Table 4).

**Table 4.**
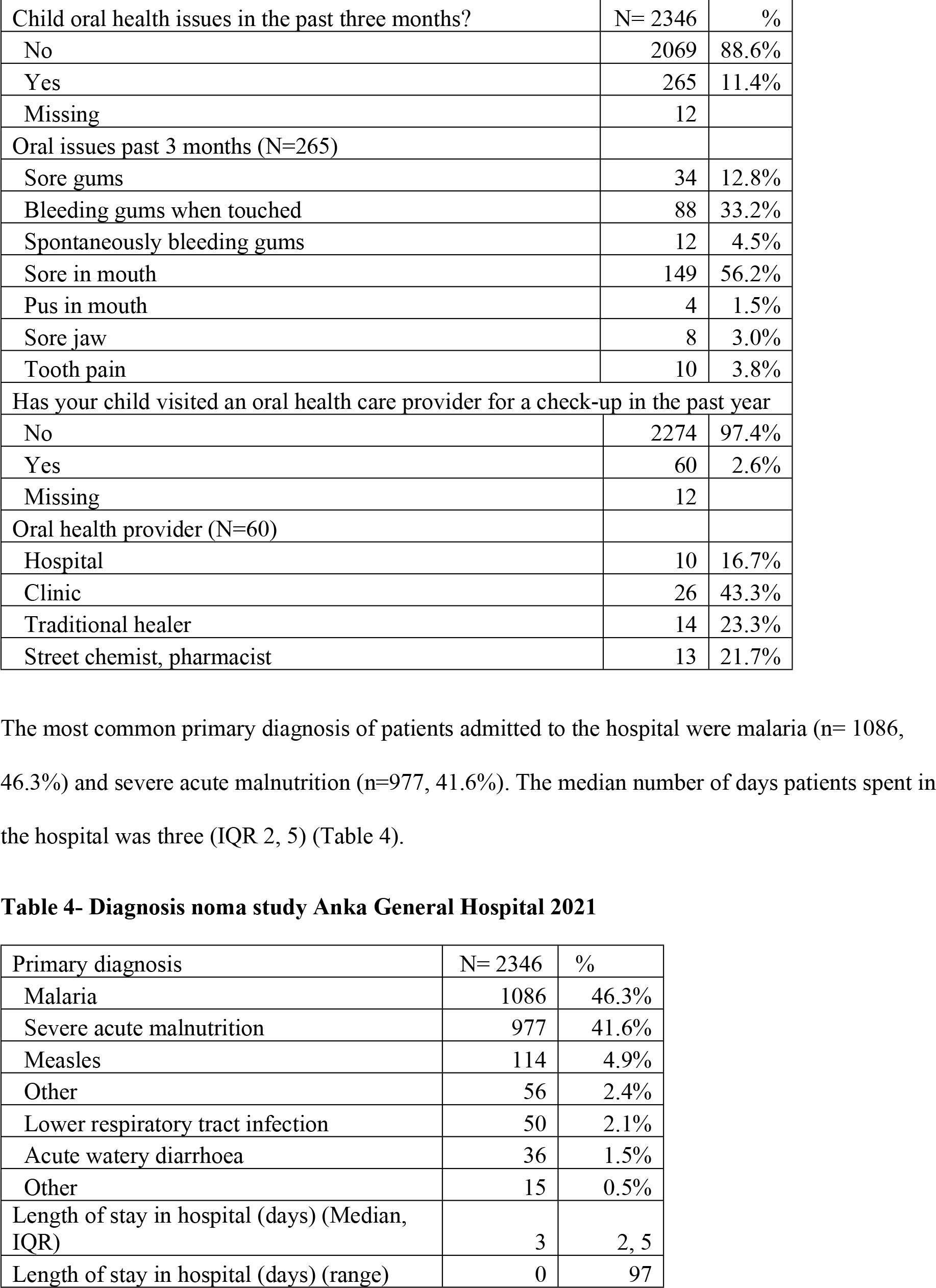
Diagnosis noma study Anka General Hospital 2021

### Gingivitis and noma diagnoses

Simple gingivitis was diagnosed in 58 patients (2.5%) and acute necrotizing gingivitis was diagnosed in six patients (0.3%) at admission. At discharge simple gingivitis was diagnosed in nine patients (0.5%) and acute necrotizing gingivitis was diagnosed in one patient (0.1%) (Table 5).

**Table 5.** Noma diagnosis upon admission and discharge, noma study Anka General Hospital 2021

| Admission Noma Stage | N= 2346 | % |
| --- | --- | --- |
| None | 2256 | 97.2% |
| Stage 0: Simple gingivitis | 58 | 2.5% |
| Stage 1: Acute necrotizing gingivitis | 6 | 0.3% |
| Missing | 26 |  |
| Discharge Noma Stage |  |  |
| None | 1667 | 99.4% |
| Stage 0: Simple gingivitis | 9 | 0.5% |
| Stage 1: Acute necrotizing gingivitis | 1 | 0.1% |
| Missing | 669 |  |

Of the 34 participants who were diagnosed with simple gingivitis on admission and who had an oral exam at discharge: 28 (82.4%) had resolved at discharge; five (14.7%) still had simple gingivitis at discharge; and one (2.9%) had acute necrotizing gingivitis upon discharge. One of the two patients who had acute necrotizing gingivitis at admission and had an oral exam at discharge, had no stage of noma at discharge, the other had simple gingivitis at discharge.

### Malnutrition

Of those patients enrolled in the study who were aged between 6 months and five years and who had a MUAC measurement completed upon admission (n=2039), n=692 (34.0%) had SAM, n=399 (19.6%) had MAM and n=1091 (53.5%) had GAM (Table 6).

**Table 6.** Malnutrition (SAM, MAM, GAM) upon admission in those aged 6 months to 5 years (n=2039) and BMI-for-age Z-scores classification for six to 12 year olds [36]

|  | TOTAL |  | ITFC |  | Not ITFC |  | Ch sq |
| --- | --- | --- | --- | --- | --- | --- | --- |
| MUAC | N=2039 | % | N=956 | % | N=1083 | % |  |
| Normal, MUAC>125mm | 948 | 46.5% | 133 | 13.9% | 815 | 75.3% | <0.05 |
| SAM, MUAC <115mm | 692 | 33.9% | 604 | 63.2% | 88 | 8.1% |  |
| MAM, between MUAC ≥115 and <125mm | 399 | 19.6% | 218 | 22.9% | 180 | 16.6% |  |
| GAM, MUAC <125mm | 1091 | 53.5% | 823 | 86.1% | 268 | 24.8% | <0.05 |
| BMI-for-age Z-score | N=100 | % | N=31 | % | N=69 | % |  |
| Severe thinness (<-3SD) | 28 | 28.0% | 16 | 51.6% | 12 | 17.4% | <0.05 |
| Thinness (≥ -3 SD & < -2 SD) | 16 | 16.0% | 2 | 6.5% | 14 | 20.3% |  |
| Normal (≥ -2 SD & < +1SD) | 51 | 51.0% | 10 | 32.3% | 41 | 59.2% |  |
| Overweight (≥+1SD & <+2 SD) | 2 | 2.0% | 1 | 3.2% | 1 | 1.5% |  |
| Obesity (≥+2SD) | 3 | 3.0% | 2 | 6.5% | 1 | 1.5% |  |

Of the participants aged between six and 12 years who had weight and height measured upon admission (n=100), 44.0% (n=44) were wasted (BMI/age <-2 z-score). The majority of those admitted to the ITFC were wasted (n=18, 58.1%), and the majority of those not admitted to the ITFC were classified with normal BMI z-scores (n-41, 59.2%) (Table 6).

### Risk factors

Being aged over two years (2-6yr old odds ratio (OR) 3.4, CI 1.77, 6.5; 7 to 12 yr OR 5.0, CI 1.7, 14.6; p=<0.001) and being admitted to the ITFC (OR 2.1; CI 1.22, 3.62) were associated with having simple gingivitis. Those whose primary diagnosis was malaria, were less likely to have simple gingivitis upon admission compared to those admitted for other reasons (OR 0.29; CI 0.15, 0.55; p=<0.001). Of those diagnosed with acute necrotizing gingivitis (n=6) n=4/6 were aged 2-6 years, were in the lower wealth score (indicating lower socioeconomic status) and ate pap; n=5/6 children had been sick in the 3 months prior to admission (Table 7).

**Table 7:**
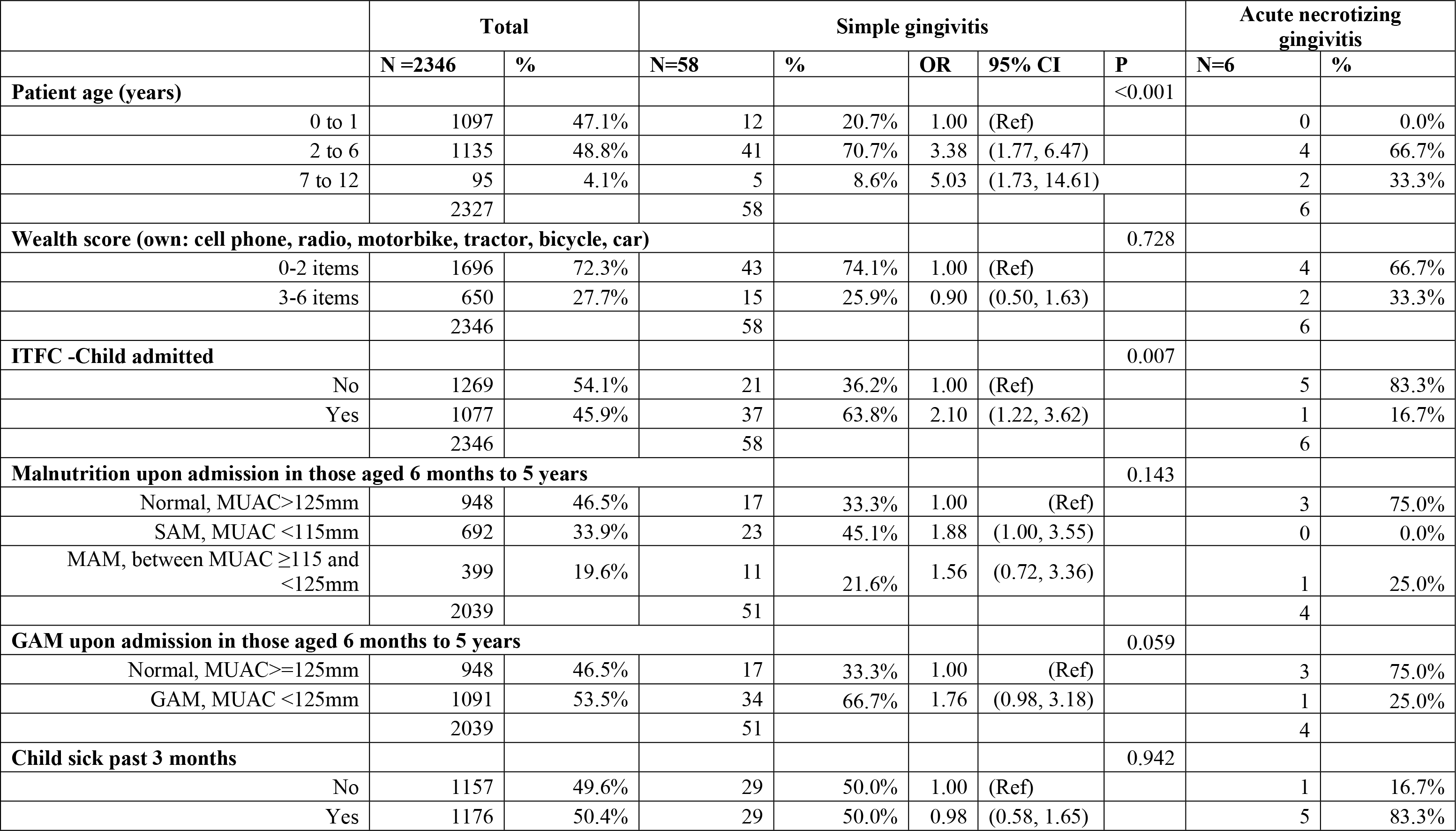
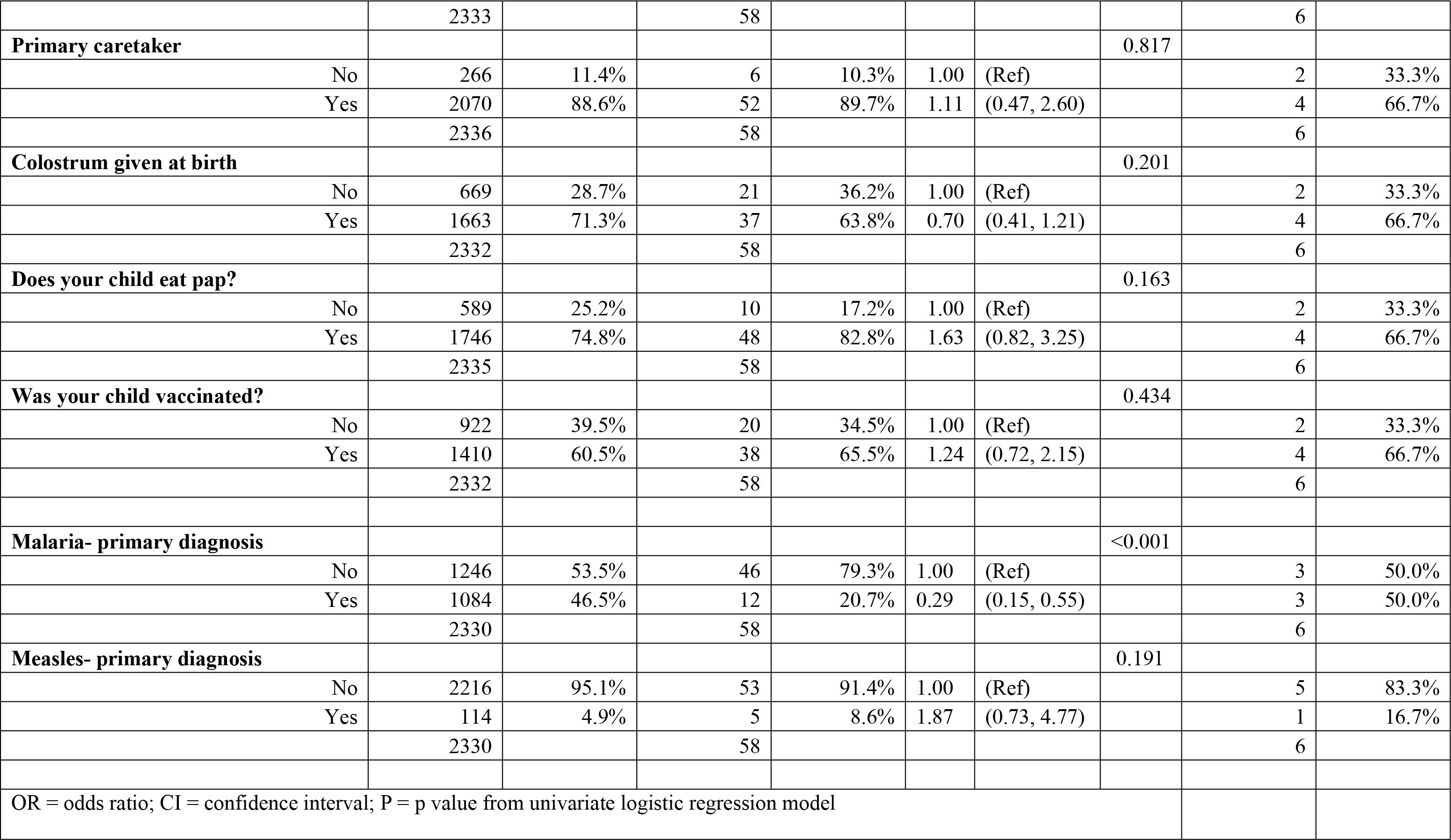
Risk factors for stage 0 and 1 on admission

|  | Total |  | Simple gingivitis |  |  |  |  | Acute necrotizing gingivitis |  |
| --- | --- | --- | --- | --- | --- | --- | --- | --- | --- |
|  | N =2346 | % | N=58 | % | OR | 95% CI | P | N=6 | % |
| **Patient age (years)** |  |  |  |  |  |  | <0.001 |  |  |
| 0 to 1 | 1097 | 47.1% | 12 | 20.7% | 1.00 | (Ref) |  | 0 | 0.0% |
| 2 to 6 | 1135 | 48.8% | 41 | 70.7% | 3.38 | (1.77, 6.47) |  | 4 | 66.7% |
| 7 to 12 | 95 | 4.1% | 5 | 8.6% | 5.03 | (1.73, 14.61) |  | 2 | 33.3% |
|  | 2327 |  | 58 |  |  |  |  | 6 |  |
| **Wealth score (own: cell phone, radio, motorbike, tractor, bicycle, car)** |  |  |  |  |  |  | 0.728 |  |  |
| 0-2 items | 1696 | 72.3% | 43 | 74.1% | 1.00 | (Ref) |  | 4 | 66.7% |
| 3-6 items | 650 | 27.7% | 15 | 25.9% | 0.90 | (0.50, 1.63) |  | 2 | 33.3% |
|  | 2346 |  | 58 |  |  |  |  | 6 |  |
| **ITFC -Child admitted** |  |  |  |  |  |  | 0.007 |  |  |
| No | 1269 | 54.1% | 21 | 36.2% | 1.00 | (Ref) |  | 5 | 83.3% |
| Yes | 1077 | 45.9% | 37 | 63.8% | 2.10 | (1.22, 3.62) |  | 1 | 16.7% |
|  | 2346 |  | 58 |  |  |  |  | 6 |  |
| **Malnutrition upon admission in those aged 6 months to 5 years** |  |  |  |  |  |  | 0.143 |  |  |
| Normal, MUAC>125mm | 948 | 46.5% | 17 | 33.3% | 1.00 | (Ref) |  | 3 | 75.0% |
| SAM, MUAC <115mm | 692 | 33.9% | 23 | 45.1% | 1.88 | (1.00, 3.55) |  | 0 | 0.0% |
| MAM, between MUAC ≥115 and <125mm | 399 | 19.6% | 11 | 21.6% | 1.56 | (0.72, 3.36) |  | 1 | 25.0% |
|  | 2039 |  | 51 |  |  |  |  | 4 |  |
| **GAM upon admission in those aged 6 months to 5 years** |  |  |  |  |  |  | 0.059 |  |  |
| Normal, MUAC>=125mm | 948 | 46.5% | 17 | 33.3% | 1.00 | (Ref) |  | 3 | 75.0% |
| GAM, MUAC <125mm | 1091 | 53.5% | 34 | 66.7% | 1.76 | (0.98, 3.18) |  | 1 | 25.0% |
|  | 2039 |  | 51 |  |  |  |  | 4 |  |
| **Child sick past 3 months** |  |  |  |  |  |  | 0.942 |  |  |
| No | 1157 | 49.6% | 29 | 50.0% | 1.00 | (Ref) |  | 1 | 16.7% |
| Yes | 1176 | 50.4% | 29 | 50.0% | 0.98 | (0.58, 1.65) |  | 5 | 83.3% |
|  | 2333 |  | 58 |  |  |  |  | 6 |  |
| **Primary caretaker** |  |  |  |  |  |  | 0.817 |  |  |
| No | 266 | 11.4% | 6 | 10.3% | 1.00 | (Ref) |  | 2 | 33.3% |
| Yes | 2070 | 88.6% | 52 | 89.7% | 1.11 | (0.47, 2.60) |  | 4 | 66.7% |
|  | 2336 |  | 58 |  |  |  |  | 6 |  |
| **Colostrum given at birth** |  |  |  |  |  |  | 0.201 |  |  |
| No | 669 | 28.7% | 21 | 36.2% | 1.00 | (Ref) |  | 2 | 33.3% |
| Yes | 1663 | 71.3% | 37 | 63.8% | 0.70 | (0.41, 1.21) |  | 4 | 66.7% |
|  | 2332 |  | 58 |  |  |  |  | 6 |  |
| **Does your child eat pap?** |  |  |  |  |  |  | 0.163 |  |  |
| No | 589 | 25.2% | 10 | 17.2% | 1.00 | (Ref) |  | 2 | 33.3% |
| Yes | 1746 | 74.8% | 48 | 82.8% | 1.63 | (0.82, 3.25) |  | 4 | 66.7% |
|  | 2335 |  | 58 |  |  |  |  | 6 |  |
| **Was your child vaccinated?** |  |  |  |  |  |  | 0.434 |  |  |
| No | 922 | 39.5% | 20 | 34.5% | 1.00 | (Ref) |  | 2 | 33.3% |
| Yes | 1410 | 60.5% | 38 | 65.5% | 1.24 | (0.72, 2.15) |  | 4 | 66.7% |
|  | 2332 |  | 58 |  |  |  |  | 6 |  |
| **Malaria- primary diagnosis** |  |  |  |  |  |  | <0.001 |  |  |
| No | 1246 | 53.5% | 46 | 79.3% | 1.00 | (Ref) |  | 3 | 50.0% |
| Yes | 1084 | 46.5% | 12 | 20.7% | 0.29 | (0.15, 0.55) |  | 3 | 50.0% |
|  | 2330 |  | 58 |  |  |  |  | 6 |  |
| **Measles- primary diagnosis** |  |  |  |  |  |  | 0.191 |  |  |
| No | 2216 | 95.1% | 53 | 91.4% | 1.00 | (Ref) |  | 5 | 83.3% |
| Yes | 114 | 4.9% | 5 | 8.6% | 1.87 | (0.73, 4.77) |  | 1 | 16.7% |
|  | 2330 |  | 58 |  |  |  |  | 6 |  |
| OR = odds ratio; CI = confidence interval; P = p value from univariate logistic regression model |  |  |  |  |  |  |  |  |  |

## Discussion

A small proportion of those enrolled in this study had simple gingivitis (2.5%) or acute necrotizing gingivitis (0.3%) at admission, and no later stages of noma were identified in the cohort. Oral health practices were not optimal and access to oral health care was limited and underutilised. Those in the ITFC were more likely to have simple gingivitis. Known risk factors for noma (malnutrition and being aged between two and six years) were associated with simple gingivitis in our cohort.

Our findings were similar to a community-based noma study in northwest Nigeria in 2020 where simple gingivitis was identified in 3.1% (n=181; 95% CI 2.6 to 3.8), acute necrotising gingivitis in 0.1% (n=10; CI 0.1 to 0.3) and oedema in 0.05% (n=3; CI 0.02 to 0.2) [34]. These similarities were expected as the studies were conducted a similar region (northwest Nigeria). However, as our study was conducted in a hospital setting with a large proportion of patients being treated for malnutrition, it could have been expected that we would have seen more noma cases being diagnosed. One plausible explanation for this not being the case, is that the age range of our survey included a significant number of children aged under two years (47.1%), which is an age group less at risk of developing noma.

A study conducted in Cameroon in 2015 including school children (n=2287) [37] found similar proportions of cases with gingivitis as in our study (2.1% had moderate gingivitis which could reflect our simple gingivitis cases and 0.6% had severe gingivitis which could reflect our acute necrotizing gingivitis cases), however comparison is difficult as the study methods and definitions differed significantly.

The caregivers in our cohort reported several challenges accessing care; these are similar to challenges reported in other noma research work in this setting, including difficulties accessing transport and the long distances patients and their caregivers need to travel in order to access care [38,39]. These issues should be taken into consideration for programs aiming to alleviate barriers to health care in this setting.

Many of the respondents who sought care sought care for a prior illness first visited a street pharmacist (39.7%) showing that they would be important agents for promoting oral health and awareness about noma. This finding is supported by another study in Northern Nigeria which showed that engaging community pharmacists may help to reduce oral health disparities by increasing oral health awareness and improving the quality of life via cost-effective delivery of pharmacy-based oral health-care services [40].

Many respondents reported cleaning their teeth (86.6%), however the main utensil used was water (84.7%), and only 10.4% reported using a toothbrush. The importance of tooth brushing and mechanisms to encourage and facilitate tooth brushing have been discussed in other literature [2,37,41–43]. In our cohort, the main reasons for children cleaning their teeth were to get clean, bright teeth (32.5%) and to get rid of foul breath (57.1%). The main reasons for children not cleaning their teeth, were not knowing the benefits of cleaning (7.2%) and forgetting to clean their teeth (7.6%).

These provide useful starting points for oral hygiene trainings which should focus on educating children and caregivers on the benefits of tooth brushing, teaching mechanisms for instilling toothbrushing into the daily routine, and explaining that toothbrushing will lead to having nice smelling breath and clean, bright teeth.

The most common oral health issues patients reported having in the prior three months were a sore in the mouth (56.2%), bleeding gums when touched (33.2%) and sore gums (12.8%). These issues are all likely varying degrees of gingivitis and show that children living in the communities served by the hospital show the clinical effects of poor oral hygiene and that most oral complaints are transient, and not chronic or progressive.

Almost all the respondents (97.4%) had not had an oral health check in the past year. This low level of access and uptake of oral care is similar to other studies in the setting [44–47]. The main reasons for the lack of oral health care that have been reported are a lack of human resources, high costs of care, sparse provision of services and low levels of knowledge about different treatment options for oral conditions [44–47]. These barriers indicate the need to incorporate oral health into primary health care in this setting, a call common in oral health literature [45–47]. In terms of noma, it would be worthwhile encouraging the inclusion of oral examinations in primary health care provision, malnutrition surveys and vaccination campaigns, particularly in vulnerable populations.

Those admitted to the ITFC were more likely to be diagnosed with simple gingivitis. This adds weight to the link between noma and malnutrition [8–10,28–30]; however, caution is needed when interpreting these findings as most cases of simple gingivitis do not lead to noma.

The most common primary diagnosis of patients admitted to the hospital were malaria (46.3%) and severe acute malnutrition (41.6%). Both of these conditions have been reported as risk factors for the development of noma [8–10,48]. However, our univariate analysis did not support this, possibly because our analysis was looking at risk factors for simple gingivitis, whereas the references are reporting risk factors for late stage noma. Another limitation is that our analysis was based on diagnosis at the same time as assessment of any stage of noma, whereas the references discuss diagnosis of comorbidities in the three months prior to onset of noma.

A further limitation was that we only recruited patients during the day time, this could have meant that patients with serious, acute complications (such as late stage noma) admitted at night time, were not included in our survey.

Our survey was conducted during the rainy season, which could have resulted in a possible bias, as this time of year could have meant increased difficulties for sicker patients to travel to the hospital which could have meant fewer later stage noma patients reached care. It could also have meant increased food insecurity, or difference in the epidemiology of other morbidities in the area. Future studies in other seasons or the year-round inclusion or oral screenings in routine activities could offer insight into whether or not the season affects the number of simple and acute necrotizing gingivitis cases identified in this context.

## Conclusion

Our study showed a small proportion of those admitted to the Anka General Hospital, Zamfara, had simple or acute necrotizing gingivitis, most of which had resolved upon discharge. Those admitted to the ITFC were more likely to have simple gingivitis, providing evidence of the link between malnutrition and the warning sign of noma. Many of those in our study cohort exhibited known risk factors for noma such as being malnourished, having a comorbidity, low socioeconomic status, poor oral hygiene practices, and a lack of access to healthcare, indicating there is a large population at risk of developing later stage noma in this setting. The lack of access to and uptake of oral health care indicates a strong need for oral exams to be included in routine health services. This provision could improve the oral status of the population and decrease the chance of patients developing later stage noma.

## Author Statements

### Acknowledgements

Thank you to the participants in this study. A big thank you to the study team for all their hard work and dedication to this project. Thanks to Vincent Kimui Ndichu, Christian Mwemezi and Rahima Shuaibu for their contribution to the study. We would also like to thank all the health care workers who are advocating for and treating noma patients around the world.

### Ethics

The MSF Ethics Review Board (ERB) (2017), Nigerian Federal Ministry of Health ERB (NHREC/01/01/2007-29/04/2021), Zamfara Ministry of Health ERB (ZSHREC01112020) and the Usman Danfodiyo University Teaching Hospital Health Research and Ethics Committee in Nigeria (NHREC/30/012/2019) approved the study protocol.

### Data Availability Statement

MSF has a managed access system for data sharing that respects MSF’s legal and ethical obligations to its patients to collect, manage and protect their data responsibility. Ethical risks include but are not limited to the nature of MSF operations and target populations being such that data collected often involves highly sensitive data. The dataset supporting the conclusions of this article is available on request in accordance with MSF’s data sharing policy (available at: http://fieldresearch.msf.org/msf/handle/10144/306501). Requests for access to data should be made to.

### Author contributions

Elise Sarah Farley (0000-0003-4457-4068): Conceptualization, Data Curation, Formal Analysis, Investigation, Methodology, Writing – Original Draft Preparation.

Miriam Njoki Karinja (0000-0003-0474-9492): Data Curation, Investigation, Project Administration, Supervision, Writing – Review & Editing.

Abdulhakeem Mohammed Lawal: Data Curation, Investigation, Project Administration, Supervision, Writing – Review & Editing.

Michael Olaleye (0009-0003-4101-8469): Data Curation, Investigation, Project Administration, Supervision, Writing – Review & Editing.

Sadiya Muhammad: Investigation, Writing – Review & Editing. Maryam Umar: Investigation, Writing – Review & Editing.

Fatima Khalid Gaya: Investigation, Writing – Review & Editing. Shirley Chioma Mbaeri: Investigation, Writing – Review & Editing.

Mark Sherlock (0009-0005-8186-4365): Conceptualization, Funding Acquisition, Writing – Review & Editing.

Deogracia Wa Kabila: Funding Acquisition, Writing – Review & Editing. Miriam Peters: Funding Acquisition, Writing – Review & Editing.

Joseph Samuel: Conceptualization, Funding Acquisition, Writing – Review & Editing. Guy Maloba: Funding Acquisition, Writing – Review & Editing.

Rabi Usman (0000-0001-7869-5357): Writing – Review & Editing.

Saskia van der Kam (0000-0002-3608-8054): Conceptualization, Writing – Review & Editing. Koert Ritmeijer (0000-0001-5924-872X): Conceptualization, Writing – Review & Editing.

Cono Ariti (0000-0001-7615-0935): Conceptualization, Formal Analysis, Methodology, Supervision, Writing – Review & Editing.

Mohana Amirtharajah (0000-0002-4779-0308): Conceptualization, Funding Acquisition, Methodology, Supervision, Writing – Review & Editing.

Grégoire Falq (0009-0002-2116-6475): Formal Analysis, Supervision, Writing – Review & Editing.

